# Sucrose-stimulated Salivary pH as an Adjunct to Caries Risk Assessment

**DOI:** 10.1101/2022.07.31.22278249

**Authors:** David Okuji, Olutayo Odusanwo, Yinxiang Wu, Susanna Yeh, Sohini Dhar

## Abstract

**Background:** Child and maternal sucrose-stimulated salivary pH (**SSS pH**) levels have the potential to be associated with childhood caries.

**Aim:** This study investigated the relationships among child and maternal SSS pH and child caries diagnosis, severity, and risk.

**Design:** SSS pH levels were measured from 202 pediatric subjects and 175 mothers. Early childhood caries (**ECC**) and severe ECC (**SECC**) diagnoses, caries risk assessment (**CRA**) results, and caries severity scores were recorded. The associations between child and maternal SSS pH and children’s caries risk, diagnosis, and severity were respectively assessed using regression models.

**Results:** Children with SSS pH ≤5.6 had higher odds to be diagnosed with ECC or SECC (aOR=7.27), and higher odds to present with moderate to extensive caries severity (aOR=5.63). Child SSS pH was associated with multiple risk factors on a CRA. When child SSS pH was adjusted for age, gender, and race/ethnicity as a predictor for SECC and ECC, the sensitivity and specificity estimates increased. Maternal and child SSS pH were positively associated.

**Conclusions:** Dentists should consider the use of children’s SSS pH as an inexpensive adjunct to the CRA and know that maternal and child SSS pH are significantly associated.

## Introduction

Dental caries is the most prevalent chronic condition in children, and if left untreated, can lead to significant comorbidities. The American Association of Pediatric Dentistry (**AAPD**) classifies early childhood caries (**ECC**) is the presence of at least one decayed, missing (due to caries), or filled tooth surfaces in a child younger than 6 years.^1^ Severe early childhood caries (**SECC**) considers disease with rampant, acute, or progressive characteristics. Conventional risk factors for ECC include low maternal education, low family socioeconomic status (**SES**), and increased consumption of refined carbohydrates.^2,3^ In recent years, unprecedented gains in the understanding of the biology and mechanism underlying human health and disease have been made, although the existing caries risk assessment and ECC prediction tools have shown limited practical clinical utility.^4^

Studies on caries risk assessment (**CRA**) show that although multiple factors influence risk, caries experience is still the strongest predictor of individual risk.^5^ However, this factor does not reach the end goal of preventing absolute caries experience, nor is it part of the causal pathway of disease. To date, it has been shown that most current CRA tools fail to consistently inform clinical decisions,^6^ however many clinicians believe CRA has a great potential to strengthen care.^7,8^

Saliva has a critical function in maintaining oral health by rapidly neutralizing the acids and promoting remineralization. In the 1940s, Dr. Robert Stephan demonstrated that following the intake of fermentable carbohydrates and in the presence of acidogenic bacteria, salivary and plaque pH levels decrease from resting pH to a pH below the critical level of 5.5, initiating the demineralization of enamel. These lower pH levels gradually return to resting pH levels and remineralization of the enamel structure takes place. Repeated intakes of fermentable carbohydrates lead to low salivary pH for extended periods, thereby preventing remineralization.^2,9,10^ Stephan reported that the resting plaque pH in patients with less severe decay was closer to neutral while those with increased caries activity had a lower resting plaque pH. Additionally, after an oral glucose rinse, patients with increased caries activity demonstrated a lower and more prolonged drop in plaque pH compared to subjects with less severe caries.^2,9,10^ Thus, further evaluation of the oral microbiome through salivary pH may be a useful addition to the multifactorial CRA models.

The relationship between salivary pH and caries incidence, prevalence, and severity in children has been evaluated by multiple researchers with contrasting results. Some studies show evidence for the correlation of salivary pH and caries susceptibility, activity, and severity in children.^11,12,13,14^ However in other studies, children with active decay did not show a relationship between caries activity and salivary acidity.^15,16,17^ According to Kutsch, salivary pH testing alone has little to offer in terms of caries risk determination and clinicians must also consider the local biofilm, chemistry, and salivary composition.^3^ On the other hand, the AAPD recommends the use of a CRA tool that captures known risk factors of caries but lacks the inclusion of saliva as a risk determinant. Current CRA methods, including the AAPD CRA tool, are mostly subjective in nature. Incorporating objective methodologies is essential to accurately identify patients at increased susceptibility to caries.^18,19,20^ A recent publication by an expert panel representing the AAPD and Dental Quality Alliance recommends improving the objectivity of existing CRA tools with the addition of items that evaluate salivary biochemistry, oral microbiome and host genome.^19^

There is demonstrated association between maternal and child cariogenic bacteria and caries experience.^21,22,23,24,25^ Vertical transmission between mothers and their children has been suggested to play a major role in the spread of organisms such as mutans streptococci and lactobacilli which cause an increase of plaque’s pH-lowering and cariogenic potential.^26^ Integrating salivary effects on caries and the vertical transmission of the oral microbiome, this study examined the association between the mother’s and the child’s salivary pH response to a sucrose challenge. To date, no studies have explored the associations with maternal and child salivary pH levels and ECC outcomes.

The hypotheses for the study were there are associations between: 1) child sucrose-stimulated salivary pH (**SSS pH**) levels and AAPD definitions of ECC and severe ECC (**SECC**) diagnoses, 2) child SSS pH levels and the merged-International Caries Detection and Assessment System (**ICDAS**) classification for caries severity, 3) child SSS pH levels and risk factors categorized as “high risk” on the AAPD CRA tool, 4) child and biologic mother SSS pH levels, and (5) that SSS pH has predictive value for childhood caries.

## Methods

### Study modeling

The study was designed as an interventional, analytical, prospective, cohort study and modeled on one of the outcomes from the classic Stephan experiment, which found that after oral exposure to simple carbohydrates, the pH level of saliva and plaque decrease and gradually return to resting pH level within thirty minutes after exposure.^2,9,10^ Children and their biologic mothers who met the study’s inclusion criteria were exposed to oral sucrose and their salivary pH was measured thirty minutes after exposure. The study was also modeled on the principles of implementation science, which is defined as “the study of methods to promote the adoption and integration of evidence-based practices, interventions and policies into routine health care and public health settings.”^27^ The study design implemented evidence-based science into clinical practice by utilizing a component of the classic Stephan experiment as a cost- and time-effective tool to assist in CRA.

### Study sample

The study was approved by the NYU Grossman School of Medicine Institutional Review Board under protocol number 18-00076. An overall convenience sample of 202 pediatric subjects and their biologic mothers were selected from the pool of consecutive patients presenting to the dental clinic between March and October 2018, who met the inclusion criteria at NYU Langone Hospitals-affiliated training sites located in Massachusetts and Tennessee. Children were included in the sample if they were 0-5 years in age, categorized as American Society of Anesthesiologists (**ASA**) Physical Status Classification System of 1 (normal, healthy patient) or 2 (patient with mild systemic disease), were a patient of record at an NYU Langone Hospitals-affiliated training site, had at least one erupted tooth, had a CRA documented using the AAPD CRA tool, and had an oral evaluation for ECC diagnosis and ICDAS classification. Of the 202 enrolled mother-child dyads, 27 mothers opted not to provide saliva samples.

### Data collection

Four resident-researchers enrolled in the NYU Langone Hospitals-Advanced Education in Pediatric Dentistry residency program performed data collection after attending a training conference to ensure one standardized protocol across two study locations. The four resident-researchers diagnosed dental caries via clinical and, if available, radiographic examinations. Following the sample selection, corresponding clinical charts were reviewed using electronic medical record software. Biologic mothers of pediatric subjects, who met the inclusion criteria, were invited to participate in the study. At the visit, mothers provided informed written consent to participate in the study. Both mother and child were instructed to not brush their teeth 12 hours prior to the sucrose challenge at the next visit. The study’s premise was that a certain amount of time was required for the biofilm to develop in order to metabolize the administered sugar.^28^ At the next visit, one packet of table sugar (sucrose) was administered to the biologic mother and child subjects by dissolving the sugar granules on the mother’s and child’s tongue, respectively, and mother and child were asked to wait approximately 30 minutes before collection of their respective saliva specimen. During the 30-minute waiting period, the mother was asked to complete a sociodemographic questionnaire. After the sucrose challenge, the biologic mother and pediatric subjects expectorated whole saliva into a cup for about 1-minute. The resident-researcher measured the pH levels by dipping pH strips into the provided saliva specimen and compared immediate color change with a chart provided by the manufacturer (Precision pH 4070 test strips, Precision Laboratories, Cottonwood, AZ) and recorded the child’s and mother’s pH level into the child subject’s electronic dental record. The pH strip color chart measured pH in eight ordinal increments at pH levels 4.0, 4.4, 4.8, 5.2, 5.6, 6.0, 6.5, and 7.0. The pH strips have a reported accuracy of ± ½ color chart unit or ± 0.2 pH units (Precision pH 4070 test strips, Precision Laboratories, Cottonwood, AZ). Clinical data from the dental record, including the child’s caries diagnosis and caries severity (using the merged-ICDAS system) were abstracted after the collection of saliva specimens.^9,29,30^ The de-identified clinical data was entered onto a paper data collection form and subsequently transferred into the password-secured web-based Research Electronic Data Capture data collection platform (REDCap®; Vanderbilt University, Nashville, TN). Data entry of study variables from the clinical charts were performed by resident-researchers from each of the participating clinic locations. Under the direction of the Principal Investigator of the study and local faculty mentors, resident-researchers ensured participant confidentiality. Data management and quality assurance were also ensured.

### Statistical analysis

For descriptive statistics, continuous variables were summarized with mean and standard deviations and categorical variables were summarized with counts and percentages. The descriptive statistics were reported overall, by child caries diagnosis and by different levels of pH of child saliva specimens. The distributions of variables were compared across different groups with the two-sample independent t-test (or the ANOVA test, as appropriate) for continuous variables and the Chi-square (or the Fisher’s Exact test, as appropriate) for categorical variables. When assessing the correlation between child and mother salivary pH, the Kendall’s tau rank correlation test was utilized. When analyzing bivariate associations with child salivary pH and fitting further regression models, the original pH levels were categorized into three levels: 5.6 or below, 6, and 6.5 or above, due to the small number of child subjects with pH = 5.2 or 7.

The association of child SSS pH was investigated with multiple dental outcomes, including child caries diagnosis, ICDAS caries classification, and the number of teeth present with greatest severity of dental caries. For child caries diagnosis, two binary outcomes, namely, SECC/ECC vs. no dental caries, and SECC vs. ECC/no dental caries, were considered. For merged-ICDAS caries classification, both ordinal outcome (4-level classification) and binary outcome, i.e., extensive decay/moderate decay vs. initial stage decay/sound enamel, were considered. The binary outcomes were analyzed with logistic regressions and the ordinal outcome was analyzed with cumulative proportional odds models. For the number of teeth with the greatest severity of dental caries, linear regression models were fitted. All models controlled for child age, gender, race/ethnicity. Adjusted odds ratios were reported from fitted logistic/cumulative odds models, and beta coefficients were reported from fitted linear regression models. pH = 6.5 or 7 was considered as the reference level. The likelihood ratio test was used to assess model goodness of fit. The associations of mother SSS pH with the child dental outcomes as listed above were also investigated.

Sensitivity and specificity were calculated based on cross-validation to assess predictability of child salivary pH for SECC/ECC. The logistic model with child age, gender, race/ethnicity, and salivary pH as predictors, was used as the main prediction model. Five-fold cross validation was conducted. The sample with complete data of all predictors and outcomes was randomly partitioned into 5 equal sized subsamples. The random sampling was done within the levels of the outcome to balance the class distribution within the splits. Of the 5 subsamples, a single subsample is retained as the validation data for testing the prediction model and the remaining 4 subsamples were used to fit the model. The cross-validation process was repeated 5 times, with each of the 5 subsamples used exactly once as the validation data. The whole 5-fold cross-validation was repeated 10 times. Sensitivity and specificity were estimated from 10*5 validation data sets were reported and the results were compared to that from the model with only child age, gender, race/ethnicity as the predictors.

Utilizing data from the Stephan data^10^, an *a priori* power analysis was conducted and estimated the sample size to obtain an 80% power level was five subjects.

All statistical analyses were performed in R version 4.0.3 for Mac OS.^31^ Since the dataset was used to test the primary research interest of associations of child salivary pH with multiple outcomes (10 statistical comparisons in total), the analysis adopted the Bonferroni correction and significance level was set at 0.005.

## Results

### Study population

The study included 202 child subjects. Table 1 presents the overall distribution of child-level demographic and clinical characteristics, and pH of both child and maternal saliva specimens. The pediatric subjects were 49% female and the average ± standard deviation (**SD**) age was 3.37 ± 1.22. The subjects were ethnically and racially 27.7% Non-Hispanic White, 24.3% Hispanic, and 38.1% Non-Hispanic Black. The majority (94.1%) of the subjects were healthy as defined by ASA classification, however, a large proportion (85.1%) were identified as at high-risk for caries, as determined by the AAPD CRA tool; nearly half (N = 87, 43.1%) were diagnosed with ECC and (N = 52, 25.7%) with SECC (Table 1). The subjects diagnosed with ECC/SECC were on average older and more Hispanic.

**Table 1.** Demographic and clinical characteristics of child subjects, overall and by child diagnosed caries status.

|  |  | Child dental caries |  |  |  |
| --- | --- | --- | --- | --- | --- |
|  | Overall | No Dental Caries | ECC | SECC | p |
| N (%) | 202 (100.0) | 63 (31.2) | 87 (43.1) | 52 (25.7) |  |
| Age (mean (SD)) | 3.37 (1.22) | 2.67 (1.18) | 3.75 (1.12) | 3.58 (1.07) | <0.001 |
| Sex = female (%) | 99 (49.0) | 36 (57.1) | 36 (41.4) | 27 (51.9) | 0.144 |
| Race/Ethnicity (%) |  |  |  |  | 0.003* |
| Non-Hispanic White | 56 (27.7) | 26 (41.3) | 20 (23.0) | 10 (19.2) |  |
| Hispanic | 49 (24.3) | 8 (12.7) | 22 (25.3) | 19 (36.5) |  |
| Non-Hispanic Black | 77 (38.1) | 26 (41.3) | 38 (43.7) | 13 (25.0) |  |
| Non-Hispanic Other/Multi-race | 3 (1.5) | 0 (0.0) | 1 (1.1) | 2 (3.8) |  |
| †N/A | 17 (8.4) | 3 (4.8) | 6 (6.9) | 8 (15.4) |  |
| Payer source (%) |  |  |  |  | 0.216* |
| Other | 4 (2.0) | 3 (4.8) | 1 (1.1) | 0 (0.0) |  |
| Federal Medicaid Program | 196 (97.0) | 59 (93.7) | 85 (97.7) | 52 (100.0) |  |
| †N/A | 2 (1.0) | 1 (1.6) | 1 (1.1) | 0 (0.0) |  |
| ASA = 2 (Patient with mild, systemic disease) (%) | 12 (5.9) | 3 (4.8) | 3 (3.4) | 6 (11.5) | 0.133 |
| Clinics (%) |  |  |  |  | <0.001 |
| Massachusetts Clinic | 35 (17.3) | 8 (12.7) | 9 (10.3) | 18 (34.6) |  |
| Tennessee Clinic 1 | 120 (59.4) | 30 (47.6) | 65 (74.7) | 25 (48.1) |  |
| Tennessee Clinic 2 | 47 (23.3) | 25 (39.7) | 13 (14.9) | 9 (17.3) |  |
| **ICDAS (%) |  |  |  |  | <0.001* |
| sound enamel | 63 (31.2) | 63 (100.0) | 0 (0.0) | 0 (0.0) |  |
| initial stage decay | 44 (21.8) | 0 (0.0) | 38 (43.7) | 6 (11.5) |  |
| moderate decay | 56 (27.7) | 0 (0.0) | 39 (44.8) | 17 (32.7) |  |
| extensive decay | 39 (19.3) | 0 (0.0) | 10 (11.5) | 29 (55.8) |  |
| Number of Childs teeth which present with the **ICDAS Classification with greatest severity of dental caries (mean (SD)) | 2.48 (3.05) | 0.38 (3.02) | 2.45 (1.63) | 5.06 (2.97) | <0.001 |
| Overall caries risk assessment = high risk (%) | 172 (85.1) | 37 (58.7) | 83 (95.4) | 52 (100.0) | <0.001 |
| pH of child sucrose-stimulated saliva specimen (mean (SD)) | 6.17 (0.42) | 6.38 (0.44) | 6.10 (0.35) | 6.03 (0.39) | <0.001* |
| pH of maternal sucrose-stimulated saliva specimen (mean (SD)) for overall N = 175 | 6.37 (0.41) | 6.39 (0.52) | 6.35 (0.34) | 6.38 (0.33) | 0.026* |

Mean (SD) were presented for continuous variables; N (%) were presented for categorical variables.
P Values correspond to the testing of differences by child caries diagnosis using the ANOVA test for continuous variables and using the Chi-square test (or Fisher exact test, indicated by \*) for categorical variables.
<sup>†</sup>N/A = Not applicable and was not considered as a categorical variable for statistical testing.

### Bivariate associations with child SSS pH

SSS pH of child saliva specimens were observed to be significantly associated with child caries status (Table 1 and Figure 1). Significant differences in mean child salivary pH by merged-ICDAS classification were also observed (Table 2, Figure 1: (B)).

**Figure 1.**
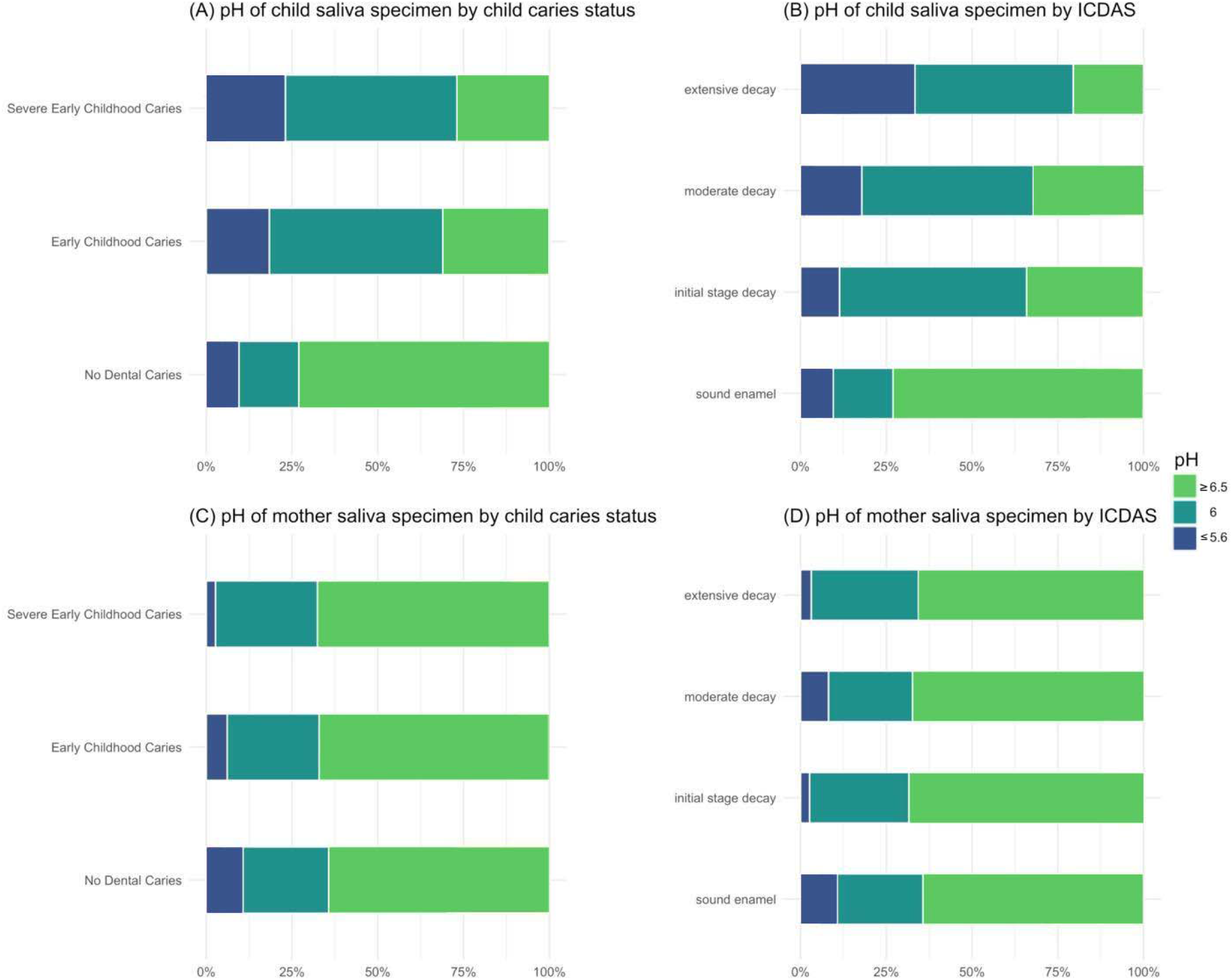
Percentage stacked bar charts by diagnosed child caries and merged-ICDAS** caries classification. (A-B): pH levels of child saliva by diagnosed child caries and merged-ICDAS caries classification; (C-D): pH levels of maternal saliva by diagnosed child caries and merged-ICDAS caries classification. Light green bars correspond to pH level = 6.5 and 7; Dark blue bars correspond to pH level = 5.2 and 5.6. ** International Caries Detection and Assessment System.^24^

**Table 2.** Demographic and clinical characteristics of child subjects, by SSS pH levels of child subjects. The saliva pH levels were grouped into ≤ 5.6, 6 and ≥ 6.5 due to the relatively small number of subjects with pH = 5.2 or 7. N = 202.

|  | pH of child saliva specimen |  |  | p-value |
| --- | --- | --- | --- | --- |
| | $\leq 5.6$ | 6 | $\geq 6.5$ | |
| N | 34 | 81 | 87 |  |
| Age (mean (SD)) | 3.56 (1.16) | 3.43 (1.14) | 3.23 (1.31) | 0.339 |
| Sex = female (%) | 20 (58.8) | 35 (43.2) | 44 (50.6) | 0.289 |
| Race/Ethnicity (%) |  |  |  | 0.328* |
| Non-Hispanic White | 9 (26.5) | 19 (23.5) | 28 (32.2) |  |
| Hispanic | 10 (29.4) | 16 (19.8) | 23 (26.4) |  |
| Non-Hispanic Black | 13 (38.2) | 36 (44.4) | 28 (32.2) |  |
| Non-Hispanic Other/Multi-race | 1 (2.9) | 2 (2.5) | 0 (0.0) |  |
| ‡N/A | 1 (2.9) | 8 (9.9) | 8 (9.2) |  |
| Payer source (%) |  |  |  | 0.523* |
| Other | 0 (0.0) | 1 (1.2) | 3 (3.4) |  |
| Federal Medicaid Program | 32 (94.1) | 80 (98.8) | 84 (96.6) |  |
| ‡N/A | 2 (5.9) | 0 (0.0) | 0 (0.0) |  |
| ASA = 2 (Patient with mild, systemic disease) (%) | 4 (11.8) | 2 (2.5) | 6 (6.9) | 0.139 |
| †ICDAS (%) |  |  |  | <0.001 |
| sound enamel | 6 (17.6) | 11 (13.6) | 46 (52.9) |  |
| initial stage decay | 5 (14.7) | 24 (29.6) | 15 (17.2) |  |
| moderate decay | 10 (29.4) | 28 (34.6) | 18 (20.7) |  |
| extensive decay | 13 (38.2) | 18 (22.2) | 8 (9.2) |  |
| Number of Childs teeth which present with the †ICDAS Classification with greatest severity of dental caries (mean (SD)) | 3.15 (2.50) | 3.25 (3.54) | 1.49 (2.44) | <0.001 |
| Overall caries risk assessment = high risk (%) | 30 (88.2) | 75 (92.6) | 67 (77.0) | 0.015 |
| Child caries diagnosis = ECC/SECC (%) | 28 (82.4) | 70 (86.4) | 41 (47.1) | <0.001 |
| pH of maternal saliva specimen (mean (SD)) | 6.11 (0.47) | 6.32 (0.34) | 6.53 (0.36) | <0.001** |

Mean (SD) were presented for continuous variables; N (%) were presented for categorical variables.
P Values correspond to the testing of differences by child caries diagnosis using the ANOVA test for continuous variables and using the Chi-square test (or Fisher exact test, indicated by \*) for categorical variables.
\*\* $p < 0.001$ from Kendall's tau rank test; Kendall's tau rank correlation = 0.33.
†International Caries Detection and Assessment System.<sup>24</sup>
‡N/A = Not applicable and was not considered as a categorical variable for statistical testing.

Table 2 presents demographic and clinical characteristics potentially associated with the pH of child saliva specimens. The original pH levels were categorized into three levels: 5.6 or below, 6, and 6.5 or above, due to the small number of child subjects with pH = 5.2 or 7. The child and maternal SSS pH were observed to be positively correlated with Kendall’s tau rank correlation = 0.33 (p<0.001), but no associations with demographics were observed. The child subjects with lower SSS pH (pH ≤ 6) had greater tendency to have overall high caries risk assessed by AAPD CRA tool, moderate/extensive decay on teeth, ECC/SECC, and more teeth with greatest severity of decay, compared to those with pH ≥ 6.5. The association with individual CRA items can be found in Figure 2.

**Figure 2.**
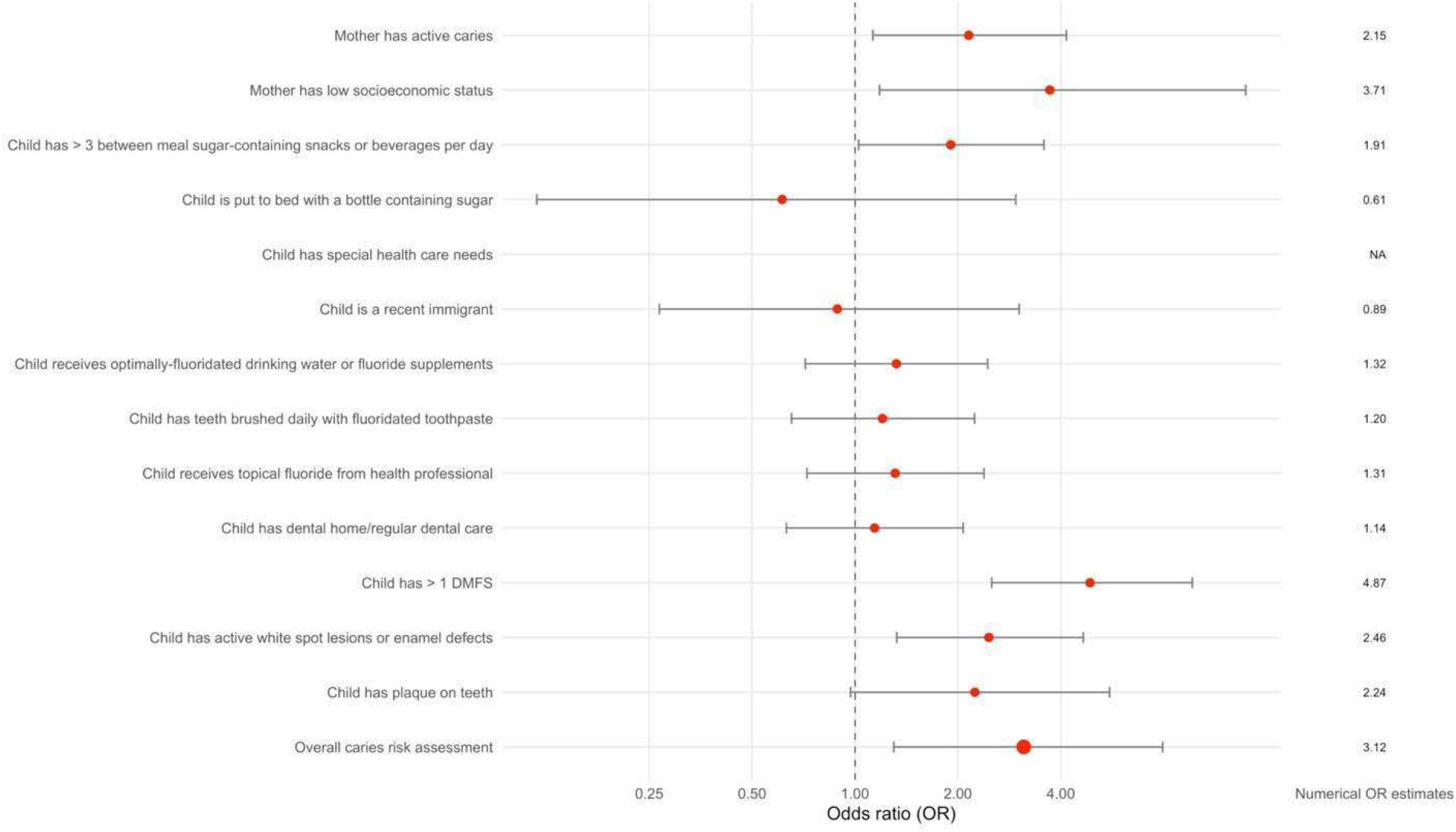
Forest plots for the associations of the pH of child saliva specimens with individual Caries Risk Assessment (CRA) factors (high risk vs. low/moderate risk). Red points correspond to unadjusted odds ratio (OR) estimates of having high risk. OR > 1 indicates that subjects with pH ≤ 6 had higher odds of being high risk for the specific risk factor. Horizontal error bars represent the 95% confidence intervals. The vertical dashed line corresponds to OR = 1. For the risk item “Child has special health care needs,” the number of subjects with high risk was below 5 and hence the OR was not present. Numerical OR estimates are provided on the right-side panel.

**Figure 3.**
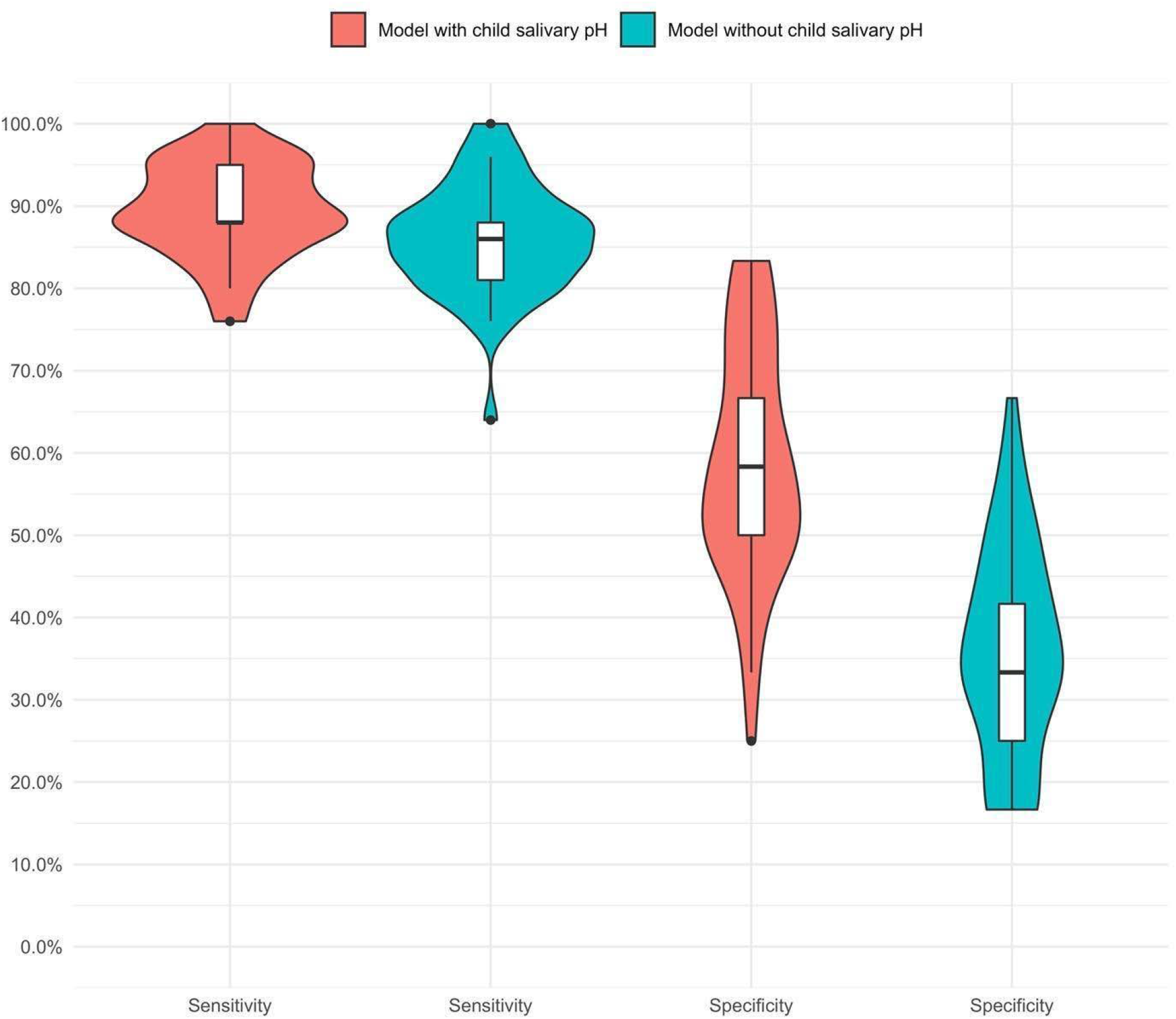
Violin plots for sensitivity and specificity estimates for predicting SECC/ECC. Red plots correspond to the logistic model with child age, gender, race/ethnicity, and salivary pH as the predictors. Blue plots correspond to the logistic model with only child age, gender, and race/ethnicity as the predictors. Sensitivity and specificity were calculated based on repeated 5-fold cross-validation to assess predictability of child salivary pH for SECC/ECC. The sample with complete data of all predictors and outcomes was N=185 and was randomly partitioned into 5 equal sized subsamples (N=37). Of the 5 subsamples, a single subsample was retained as the validation data for testing the prediction model and the remaining 4 subsamples were used to fit the model. The cross-validation process was repeated 5 times, with each of the 5 subsamples used exactly once as the validation data. The random sampling was done within the levels of the outcome to balance the class distribution within the splits. The entire 5-fold cross-validation was repeated 10 times. Sensitivity and specificity of predicting SECC/ECC over 50 (10*5) validation data sets are reported.

The adjusted associations of the child or maternal SSS pH with the dental outcomes of caries diagnosis, severity of decay, and the number of teeth with the greatest severity of decay can be found in Table 3. Statistically significant associations of child salivary pH were observed with all dental outcomes. Compared to those with salivary pH ≥ 6.5, child subjects with pH ≤ 6 had greater odds of having SECC/ECC, more severe decay on teeth, and had an increased number of teeth with the greatest severity of decay (Table 3). For example, the odds of having SECC/ECC for child subjects with salivary pH = 6 and pH ≤ 5.6 were respectively 11.11 [4.55, 29.85] and 7.27 [2.44, 24.96] times higher than that for those with salivary pH ≥ 6.5. With adjustment for child age, gender, and race/ethnicity, maternal salivary pH was not associated with any dental outcome (p-values > 0.05 from the likelihood ratio test for all fitted models).

**Table 3.** Associations of SSS pH levels with different dental outcomes. Results from logistic, cumulative odds, and regular linear models. Associations of child and maternal SSS pH with different outcomes were estimated in separate models. Complete data N = 185 when pH of child saliva was used as the predictor due to missingness in race/ethnicity. Complete data N = 158 when pH of maternal saliva was used as the predictor due to missingness in race/ethnicity and maternal saliva.

| Outcome |  |  | pH of child sucrose-stimulated saliva specimen as predictor |  | pH of maternal sucrose-stimulated saliva specimen as predictor |  |
| --- | --- | --- | --- | --- | --- | --- |
|  |  |  | aOR [95% C.I.] | p-value | aOR [95% C.I.] | p-value |
| *Child caries status* | M1: ECC/SECC vs. no caries | pH = 6 vs. ref | 11.11 [4.55, 29.85] | <0.001 | 1.41 [0.62, 3.31] | 0.419 |
|  |  | pH ≤ 5.6 vs. ref | 7.27 [2.44, 24.96] | 0.001 | 0.88 [0.22, 3.42] | 0.848 |
|  | M2: SECC vs. ECC/no caries | pH = 6 vs. ref | 3.76 [1.61, 9.38] | 0.003 | 1.44 [0.57, 3.47] | 0.425 |
|  |  | pH ≤ 5.6 vs. ref | 3.50 [1.26, 9.92] | 0.017 | 0.42 [0.02, 2.47] | 0.422 |
| *Merged-ICDAS\*\** | M3: 4-level classification (ordinal) | pH = 6 vs. ref | 5.21 [2.79, 9.95] | <0.001 | 1.26 [0.65, 2.47] | 0.494 |
|  |  | pH ≤ 5.6 vs. ref | 7.20 [3.18, 16.77] | <0.001 | 0.96 [0.29, 2.98] | 0.946 |
|  | M4: Extensive decay/moderate decay vs. initial stage decay/sound enamel | pH = 6 vs. ref | 4.16 [2.02, 8.88] | <0.001 | 1.11 [0.52, 2.35] | 0.779 |
|  |  | pH ≤ 5.6 vs. ref | 5.63 [2.28, 14.69] | <0.001 | 1.19 [0.31, 4.34] | 0.787 |
|  |  |  | Beta [95% C.I.] | p-value | Beta [95% C.I.] | p-value |
|  | M5: Number of child teeth present with greatest severity of dental caries (continuous) | pH = 6 vs. ref | 1.85 [0.93, 2.77] | <0.001 | 0.96 [-0.14, 2.05] | 0.085 |
|  |  | pH ≤ 5.6 vs. ref | 1.58 [0.41, 2.74] | 0.008 | 0.02 [-1.85, 1.89] | 0.984 |

Ref: pH = 6.5 or 7; aOR: adjusted odds ratio; C.I.: confidence intervals.
All models controlled for child age, gender, and race/ethnicity.
M1, M2, M4: Logistic regressions.
M3: cumulative odds model.
M5: linear model.
\*\* International Caries Detection and Assessment System. <sup>24</sup>

### Estimates for predicting SECC/ECC

When child age, gender, race/ethnicity, and salivary pH were used as the predictors for SECC/ECC, the median [IQR: interquartile range] of sensitivity and specificity estimates from repeated 5-fold cross-validation were respectively 88.0% [88.0%, 95.0%] and 58.3% [50.0%, 66.7%] (Figure 4). This is compared to 86.0% [81.0%, 88.0%] and 33.3% [25.0%, 41.7%] from the model with only child age, gender, race/ethnicity as the predictors.

## Discussion

The results of the study substantiate its hypotheses of associations between child SSS pH levels and AAPD definitions of ECC and SECC; ICDAS classifications; and multiple items on the AAPD CRA tool, which importantly includes classification as “high risk” for “overall caries-risk assessment.” There was no conclusive evidence to support the associations between maternal SSS pH level and these outcomes.

This study’s result of significant associations between children’s SSS pH and dental caries diagnosis and ICDAS classification aligns with previous studies. In 2013, Cunha identified mean dental caries overall was 60% higher for participants with a resting salivary pH ≤ 6.0 compared with that in participants with a resting salivary pH ≥ 6.4.^32^ A Polish study, which found salivary pH to be highly correlated with active carious lesions in children ages 7-10 years and suggested salivary pH can be used as a biomarker for predicting caries experience.^33^ There is great value in saliva being a diagnostic biomarker in caries prediction and certain assays are being developed with prognostic value for risk assessment.^34, 35^ These assays are expensive and test for caries-specific bacterial load, bacteria-derived oligosaccharides.^36,37^ However, pH test strips utilized in this study were inexpensive, costing about 20 cents per strip.

As well, this study’s result of associations between children’s SSS pH and multiple items on the AAPD CRA tool, especially with “overall assessment of the child’s dental caries risk” classified as “high,” are consistent with Gao’s similar study in 2010, who determined plaque acidity in children was associated with increased caries risk on the basis of factors such as “frequent sweet intakes” and “past caries experience.”^38^ Contrarily, a 2015 systematic review conducted by Fontana indicated salivary markers not helpful in formal CRA for children less than 5 years old, as it has been reported that a low stimulated saliva flow rate is associated with increased caries experience in adults but not in children.^39,40,41^

It has been discussed that many caries risk prediction models are inconsistently measured and reported, and consist of questionable predictors.^42^ SSS pH, which acts as a functional readout of the oral microbiome, contributes to the identification of the etiology of an individual’s caries risk. Because this study has shown that child SSS pH is a sensitive and specific predictor for SECC/ECC, the utilization of SSS pH in the caries risk model may increase consistency of the CRA and further facilitate evidence-based clinical decisions.

The strengths of this study include geographic and racial diversity of the sample population, with representation by two regions of the United States, nearly equal distribution between White and African American subjects, and an estimated 98.0% post-hoc power analysis when analyzing a diagnosis of ECC/SECC by child sucrose-stimulated salivary pH ≤6.5.

This study’s limitations include the lack of inter-rater reliability assessment for the resident-researchers; low specificity (i.e., a high rate of false positives); low sample size to support cross-validation models to further subset the dataset, with potential unmeasured or unadjusted confounding variables; and nearly 100% of the sample with low SES. The high rate of false positives may be due to confounding variables, which were not included in the study, of subject-specific protective factors for salivary function, such as buffering capacity, viscosity, flow rate, fluoride concentration; enamel strength, such as hardness, density, and fluorapatite concentration; and oral microbiome composition. The results may not be generalizable to the entire population, since children of low SES families have significant differences in oral health outcomes compared to those of moderate and high SES families.^43^

Future research might include a prospective study design with a larger and more SES-diverse sample, inclusion of additional subject-specific salivary protective factor-variables, longitudinal design, and inter-rater reliability assessment to study the predictive strength of pH screening. In addition, external validation data analysis and a broader diversity of SES levels would improve the future study designs to better analyze salivary pH as a predictor for ECC/SECC in children. As well, future studies might continue to include implementation science models for use in clinical practice.

The clinical application of this present study is to include SSS pH to augment the CRA to better identify at-risk children. In addition, SSS pH may help identify early intervention of prevention measures such motivational interviewing, chewing and swishing with antacids, and changing the oral microbiota by rinsing with plain water, chlorhexidine, or fluoride mouth rinses which may alter salivary pH and thus reduce the rate of caries development.^44,45^

## Conclusion

Based on the results of the study, the following conclusions can be made:

1. Dentists should consider the use of children’s SSS pH as an inexpensive adjunct to the CRA.
2. Maternal and child SSS pH are significantly associated.

### Bullet points

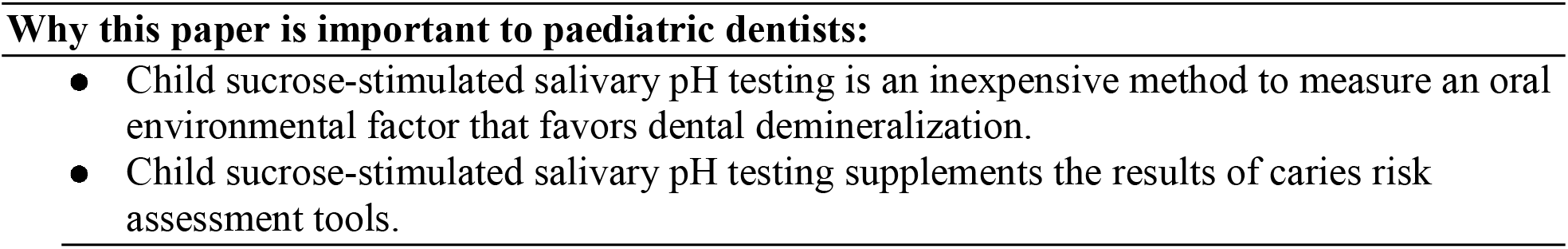

## Data Availability

All data produced in the present study are available upon reasonable request to the authors.

## Acknowledgement

The authors extend sincere appreciation and acknowledgment to dental public health specialists Martin MacIntyre, DDS, MPH and Jay Balzer, DMD, MPH; and the former resident-researchers with NYU Langone Hospitals-Advanced Education in Pediatric Dentistry: Alexander Akbari, DDS, Anita Galloway, DDS, and Gregory Poindexter, DDS.

